# Timing of the fourth dose of RTS,S/AS01 malaria vaccine in perennial settings: a modelling study

**DOI:** 10.1101/2025.10.13.25337955

**Authors:** Daniella Figueroa-Downing, Sherrie L Kelly, Aurélien Cavelan, Melissa A Penny, Josephine Malinga

## Abstract

**Background:** The recommendation of the RTS,S/AS01 malaria vaccine for use in children in moderate to high transmission areas by the WHO in 2021 provided another crucial tool in reducing malaria morbidity and mortality. As countries introduce the RTS,S vaccine, there is an interest in exploring vaccination schedules that align with existing routine health touch points and to optimize vaccine coverage.

**Methods:** This study used OpenMalaria, an individual-based, stochastic model of malaria transmission and disease progression, to assess the impact of different vaccination timing schedules and coverage on uncomplicated malaria cases, severe malaria cases, and malaria-related deaths across different archetypal transmission settings. We ran simulations comparing fourth dose protection against infection and timings at six-, nine-, 12-, 15- and 18-month intervals following the primary series, including exploring ranges of plausible protection assumptions for many of the dose timings.

**Results:** The primary series substantially reduces the malaria burden across transmission settings, regardless of timing of the fourth dose. While the number of cases averted varies by baseline prevalence, our modelling suggests that the primary series is responsible for around 54-93% of the total clinical and severe cases averted across transmission archetypes in perennial settings. Additionally, a fourth dose of the vaccine could avert additional 9-46% malaria cases and 7-41% severe cases in addition to the primary series alone. We find potential flexibility in timing this dose, especially when focused on 6 to 12 months following the primary series.

**Conclusions:** In addition to the primary series, use of a fourth dose of the RTS,S vaccine can be tailored to country specific context and linked to existing health touch points to ensure adequate coverage of this dose. In particular, a fourth dose delivered between 15- and 21-months of age with high population coverage will likely avert the largest proportion of cases of clinical malaria, severe malaria, and deaths in perennial settings, across transmission intensities.

**Trial registration:** *N/A*

## Background

Vaccines are one of the most cost-effective public health interventions for reducing morbidity and mortality in children under five, especially in low- and middle-income countries (LMICs) (1). Since its launch in 1974, vaccination through the Expanded Programme on Immunization (EPI) has helped avert an estimated 146 million deaths in children under 5 years old due to vaccine-preventable diseases, accounting for around 52% of the decline in infant mortality across the African region (2). Notably, this estimate does not include the impact of the two malaria vaccines -RTS,S/AS01 and R21/Matrix-M - recommended in 2021 and 2023, respectively for use in children in moderate and high transmission settings () (3,4). The effectiveness of immunization as a public health tool, depends not only on the availability of efficacious vaccines but also on the successful and timely delivery of all required doses. Optimizing the balance between immunological efficacy, appropriate dosing intervals, and vaccine coverage is therefore critical to achieving the full impact of malaria vaccination programs.

RTS,S is a pre-erythrocytic vaccine: a partially protective, infection blocking vaccine that induces an immune response against the circumsporozoite protein (CSP) of *Plasmodium falciparum* parasite(5– 7). In Phase 3 clinical trials, RTS,S was administered in a 4-dose schedule to 5-17 month olds with the second, third and fourth doses delivered two, three and 20 months after the first dose, respectively (8,9). The trials that concluded in 2014 showed a vaccine efficacy against clinical malaria of 36.3% (95% CI 31.8 – 40.5) and efficacy against severe malaria of 32.2% (95% CI 13.7 – 46.9) in children 5-17 months over 48 months of follow up (8). However, there was, and still is, uncertainty around duration of protection, ideal deployment schedules for maximum health impact, feasibility of reaching the targeted children and population-level impact (10,11).

Building on the Phase 3 clinical trial data, a 2015 multi-model assessment of projected public health impact and cost effectiveness estimated that a four-dose schedule of RTS,S, deployed in settings with a *Plasmodium falciparum* prevalence in 2-10 year olds (*Pf*PR_2-10_) between 10%-65% would avert a median of 116,480 (range 31,450-160,410) clinical cases and 484 (189-859) deaths per 100,000 fully vaccinated children. At US$10 per dose, close to the current market price of 9.30 euros (12), a four-dose schedule was predicted to cost US$51 (range $28-$437) per clinical case averted and $154 ($99-487) per death averted (13). These models assumed vaccine deployment at five, seven, nine and 27 months of age, corresponding to an 18-month interval between doses three and four (13). This led to the 2016 recommendation by the World Health Organization for pilot implementation studies in countries to address remaining questions around feasibility and impact prior to wide-scale introduction.

The recently concluded Malaria Vaccine Implementation Project (MVIP) pilots in Ghana, Kenya and Malawi have provided critical evidence on the implementation realities of introducing a new vaccination schedule into the existing Expanded Programme on Immunization (EPI) (14–19). The three countries chose deployment schedules that aligned to their individual country contexts with the primary series delivered between 5-9 months of age, and a fourth dose at either 22 or 24 months of age. Over the course of the pilots, coverage of the primary series reached between 60-70% of all eligible children in the three sites; however, fourth dose reach was only around 40% in all three original pilots (15,20). Despite this, all three countries demonstrated, in a real-world pilot rollout, a 13% reduction in all-cause mortality (8) following the introduction. As early evidence around the implementation challenges of introducing a novel malaria vaccine are surfaced (15,18,19,21,22), there is a critical gap that additional modelling can fill to help guide global decision making on when and how to best deploy vaccine doses, and particularly a fourth dose of malaria vaccine.

As countries in sub-Saharan Africa continue to introduce one of the two, licensed malaria vaccines, there is a continued interest in exploring dose schedules aligned to existing Expanded Programme on Immunization (EPI) touch points to optimize vaccination coverage. For example, Ghana ultimately moved their fourth dose to 18-months of age to be given alongside the second dose of measles vaccine after early pilot programs showed high dropout for a fourth dose delivered beyond the existing EPI schedule (16). After this change, the coverage of the fourth dose in Ghana rose from 42% (20) to 81% as of 2022 (23).

In this study, we provide model-based evidence for decisions around timing and deployment of a fourth dose of RTS,S in perennial settings, using age-based vaccination strategies. In parallel, another modelling group explore a similar question around dose timing in seasonal settings (24). We use mathematical modelling to estimate impact of a range of timings of the fourth dose of RTS,S vaccine on malaria morbidity and mortality in children under five years of age in in settings with varying transmission intensity. We use modelling to outline considerations for maximizing the impact of timing of the fourth dose based on both coverage and immunological response, for different settings in LMICs.

## Methods

### Transmission Model

We used OpenMalaria (https://github.com/SwissTPH/openmalaria/wiki), an individual-based, stochastic model of malaria epidemiology and control. The model simulates mosquito dynamics, vector infectiousness, blood-stage parasite densities, dynamics of infection to humans, human health dynamics to estimate incidence of malaria morbidity including severe disease and hospitalizations, and mortality (25,26). Originally developed to evaluate the population-level health impact of the RTS,S malaria vaccine (13,27), In OpenMalaria, the deployment of a pre-erythrocytic vaccine like RTS,S prevents sporozoites and therefore, infection. Additional model components and assumptions are detailed in Supplemental Table S1. The model was calibrated to uncomplicated malaria cases counts from the RTS,S Phase 3 clinical trial to derive estimates of initial protection against infection after the three-dose primary series, duration of protection (half-life, or the time it takes the initial protection against infection to reduce by 50%), and subsequent protection provided by a fourth dose (11).

### Settings and simulation scenarios

To align with previous modelling used to support WHO malaria vaccine policy, including the 2016 WHO Position Paper on Malaria Vaccines (28), we simulated archetypal transmission settings to provide the full range of probable scenarios and outcomes represented across sub-Saharan Africa (13). A factorial design was used to assess the impact of a fourth dose of RTS,S in an archetypal perennial setting. Simulated parameters include varying malaria transmission intensity (*Pf*PR_2–10_: 10%–60%, access to treatment for symptomatic cases (45%, 70%), and immunization coverage, comprising around 300 scenarios, which are described in Table 1. Simulations were run in a population of 50,000 individuals, assuming constant population growth and demography, no scale-up of vaccination coverage over time, and year-on-year immunization rates were held constant. We assumed no change in the level of other interventions during the immunization implementation period. We report model results for a range of transmission intensities, namely *Pf*PR in 2-10 year olds, ranging between 10% and 60%.

**Table 1.**
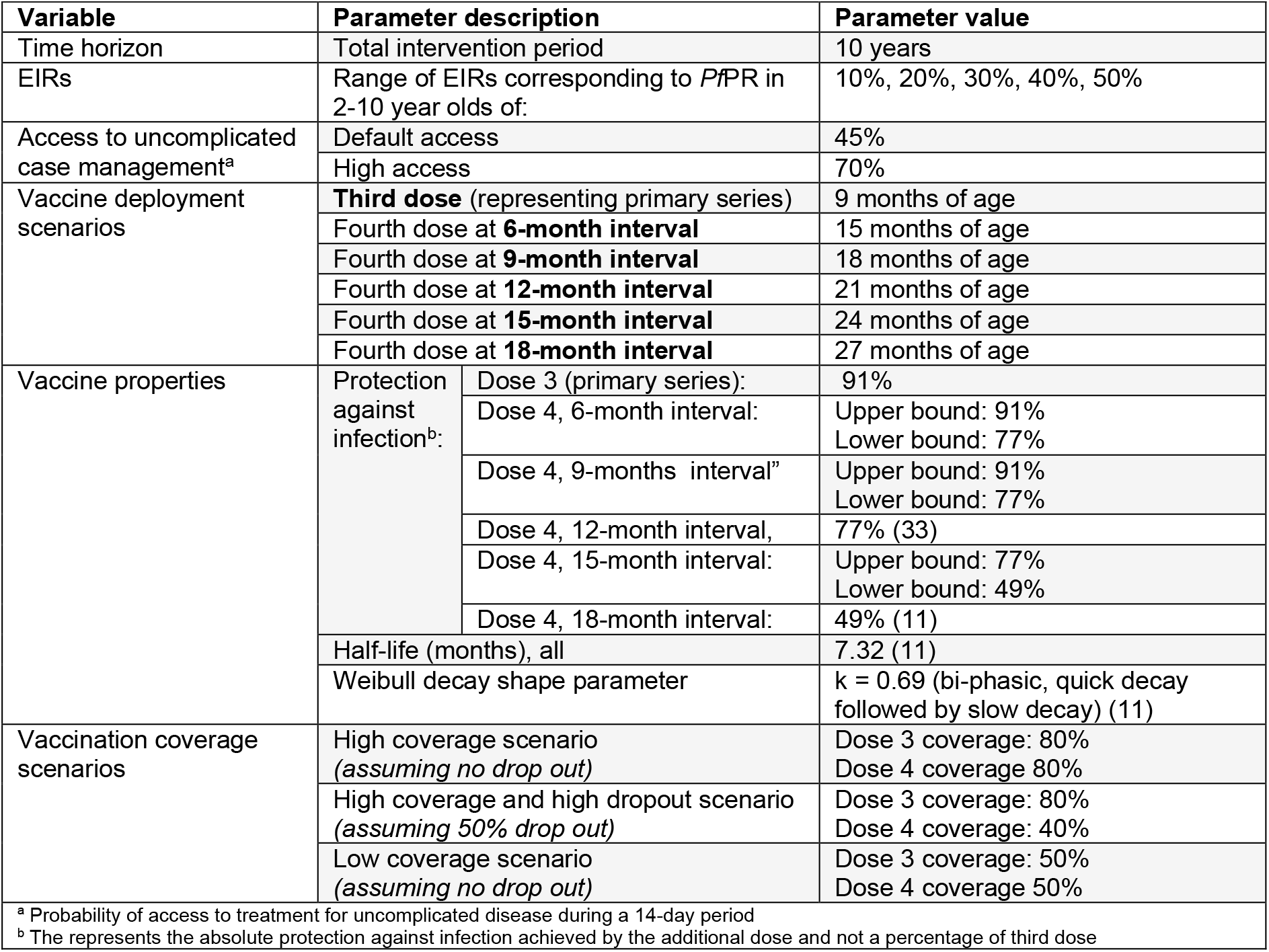
Summary of simulations: variables and levels.

### Vaccine deployment scenarios

We modelled a three-dose, age-based primary series of RTS,S with the third dose delivered at 9 months of age, in line with immunization schedules used during the MVIP pilots (20) and earlier studies (11). A fourth age-based dose was evaluated at multiple time points in the second year of life - specifically at 15, 18, 21, 24, and 27 months - representing intervals of 6 to 18 months after the primary series. These time points were selected based on stakeholder consultations and aligned with existing routine childhood vaccination visits in the first three years of life. These dose timings correspond to select countries delivering meningitis A vaccine at 15 or 18 months old (29); second dose of measles vaccine at 18 months old (30); 21 months of age to roughly reproduce Malawi’s schedule in the MVIP pilots (in Malawi the fourth dose was given at 22 months of age) (20,31); Kenya’s schedule in the MVIP pilot at 24 months (20), and at 27 months to reflect the original modelled impact from the Phase 3 clinical trials (13). We did not consider seasonal settings where additional doses (five or more doses) are recommended (3).

### Vaccine coverage and protection against infection

To examine potential differences in the impact of dose timing by vaccine coverage, we simulated three coverage scenarios: (a) a high coverage scenario with the primary series coverage of 80% and no dropout between the primary series and fourth dose (fourth dose coverage of 80%); (b) a high coverage but high dropout scenario with the primary series coverage of 80% and a 50% dropout between the primary series and fourth dose (fourth dose coverage of 40%, or roughly what was measured during early MVIP pilots); (c) a low coverage scenario with the primary series coverage of 50% and no dropout between the primary series (fourth dose coverage of 50%).

We assumed that protection against infection begin after the third dose of the primary series based on the Phase 3 trial data (8) and aligned with previous modelling work using OpenMalaria and other malaria vaccine models (11,13,32). This was estimated at 91% based on calibration of our model to the RTS,S Phase 3 clinical trial data (Table 1) (11). We assumed protection decays over time following a biphasic pattern (shape parameter k = 0.69), representing rapid initial waning followed by a slower long-term decay (11). The same decay pattern is assumed for the fourth dose, regardless of dose timing.

Finally, for the fourth dose, we based protection against infection assumptions on existing data, and logical bounds. The protection against infection for a fourth dose delivered at an 18-month interval after the primary series was based on the calibration to Phase 3 clinical trial data by Penny et al. (11). The protection against infection of a fourth dose delivered at a 12-month interval after the primary series is based on recent work on calibrating clinical trial data to OpenMalaria by Malinga et al (33) from recent field trials delivering RTS,S with and without SMC (10,34). Without existing data on efficacy assumptions for doses delivered at six-, nine-, and 15-month intervals, we explored the impact of a low and high protection against infection scenario. This range was defined by the upper and lower protection estimates of surrounding dose timings, based on previous modelling (11,33).

### Outcome measures

Public health impact was calculated as cumulative events averted per 100,000 fully vaccinated children, and age-based incidence for each outcome over a 10-year time period. We examined the impact of each of the fourth dose timings on clinical cases, severe cases, direct malaria deaths, and all malaria deaths (including those where malaria infection contributes to the outcome, in the presence of other co-occurring conditions (5,20)). Results were presented by age and across a range of *Pf*PR_2-10_, for children under 5 years of age. ‘Fully vaccinated children’ is defined as the number of children receiving at least the three-dose primary series of RTS,S vaccine. The proportion of additional cases averted by dose strategy was calculated as the proportion of cases averted by the fourth dose compared to cases averted by the primary series alone under each given scenario. All scenarios were compared against a baseline, counterfactual with no malaria vaccine intervention. Impact estimates were computed as median estimates across 10 stochastic realisations.

## Results

The three-dose primary series substantially reduced the malaria burden across all transmission intensities, regardless of timing of the fourth dose (Figure 1). In the high coverage scenario, with primary series coverage of 80% and no dropout between the third and fourth dose, the primary series accounted for approximately 54%-93% of total clinical and severe cases averted in children under five years of age across transmission settings (Table Supplement 2).

**Figure 1.**
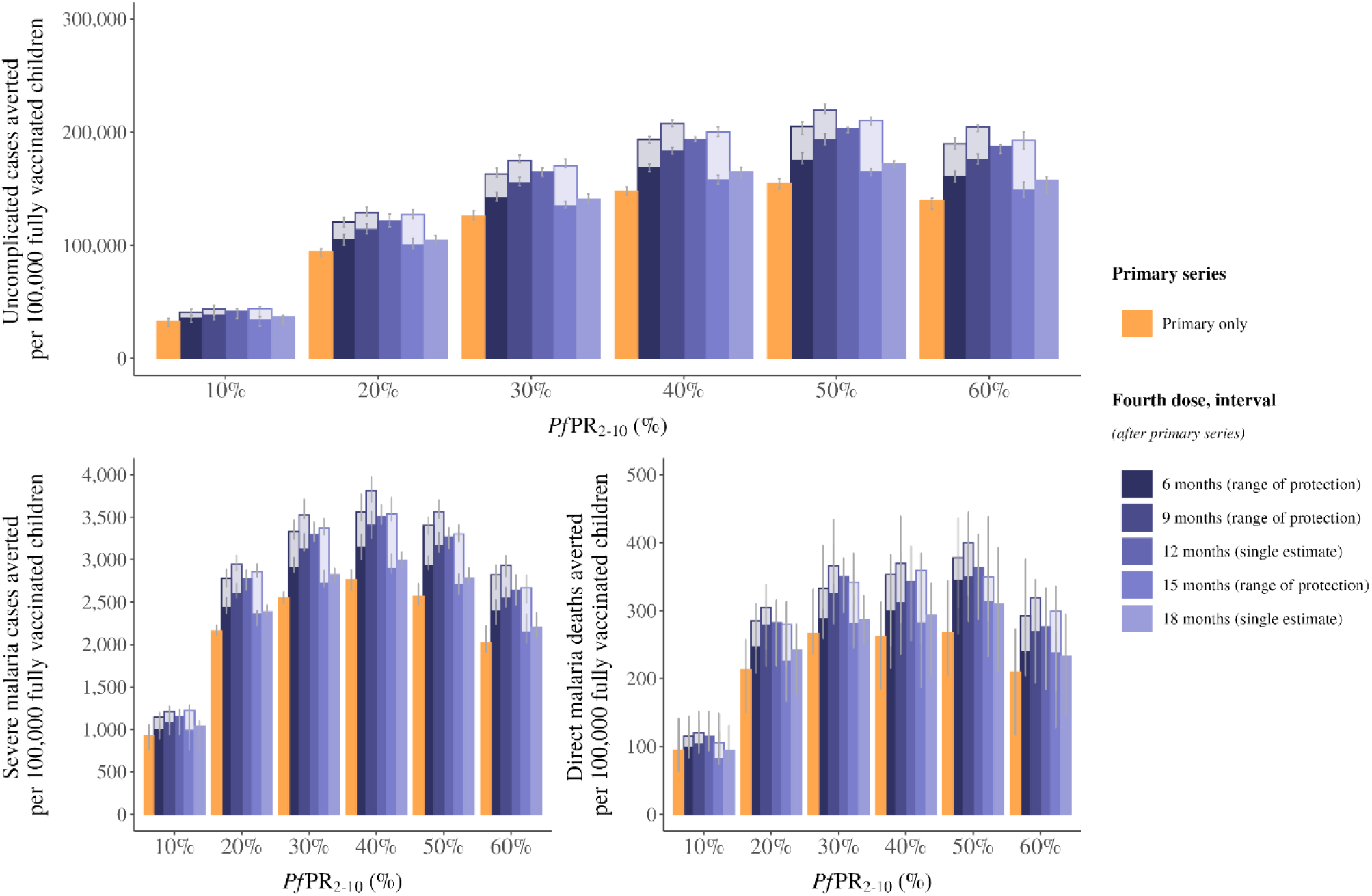
Events averted per 100,000 fully vaccinated children for fourth dose RTS,S malaria vaccine timings. The median uncomplicated malaria cases averted (A), severe malaria cases averted (B), and direct malaria deaths averted (C) in children under 5 years of age per 100,000 fully vaccinated children comparing the three-dose primary series and a fourth dose delivered at a 6-, 9-, 12-, 15- or 18-month interval after the primary series. The two shades on the 6-, 9-, and 15-month intervals represent the range of probable protection against infection values for these three dose timings. Cases averted are calculated over a 10-year time horizon. This is a default reference scenario of high coverage, where primary series coverage is assumed at 80% of the target population and there is no drop-out between the primary series and the fourth dose. Error bars show 95% confidence interval across stochastic replicates. Access to care is represented as a 45% 14-day probability to received treatment for uncomplicated malaria.

While the primary series is responsible for the majority of the impact in an age-based deployment of RTS,S, our modelling shows that a deployment of a fourth dose, especially between six and 12 months after the primary series, would likely contribute to a substantial number of additional cases averted (**Figure 2**). Across all transmission settings, a fourth dose delivered six, nine or 12 months after the primary series provide the greatest impact, though the absolute benefit varies depending on the underlying protection against infection of the dose. As we do not have empirical evidence on how the fourth dose efficacy is impacted by dose spacing, we present a plausible range of efficacies that suggest that the impact of any dose in the six to 12 month interval would likely be similar.

**Figure 2.**
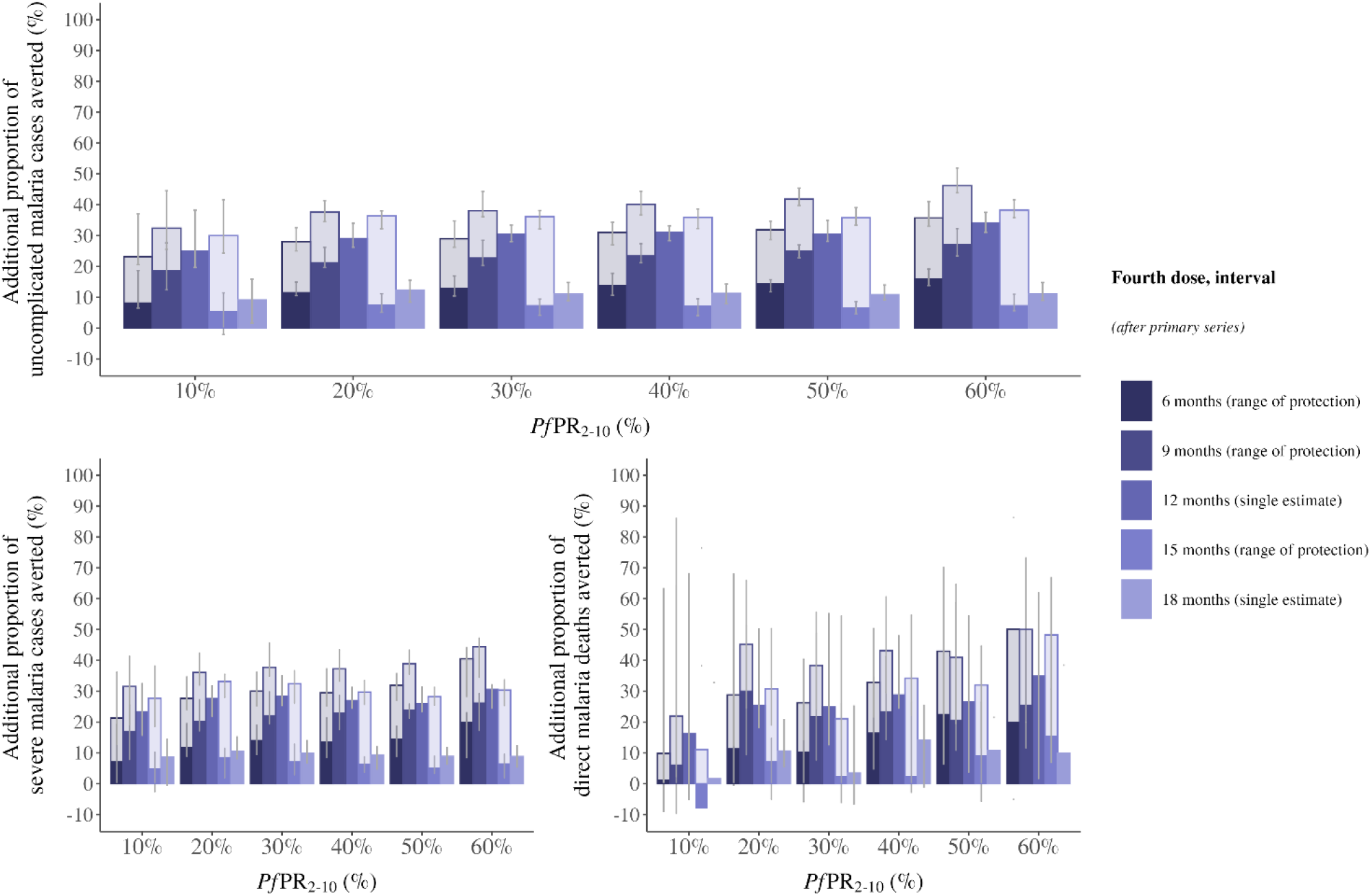
Proportion of additional cases averted by a fourth dose of RTS,S malaria vaccine. The proportion of additional uncomplicated malaria cases averted (A), severe malaria cases averted (B), and direct malaria deaths averted (C) in children under 5 years of age per 100,000 fully vaccinated children at different timings of a fourth dose delivered at a 6-, 9-, 12-, 15- or 18-month interval after the primary series, compared to the primary series alone. The two shades on the 6-, 9-, and 15-month intervals represent the range of probable protection against infection values for these three dose timings. Cases averted are calculated over a 10-year time horizon. This is a default reference scenario of high coverage, where primary series coverage is assumed at 80% of the target population and there is no drop-out between the primary series and the fourth dose. Error bars show 95% confidence interval across stochastic replicates. Access to care is represented as a 45% 14-day probability to received treatment for uncomplicated malaria.

In moderate to high transmission intensities (*Pf*PR_2-10_ > 20%), we estimate a fourth dose delivered between six and 18 months could avert an additional 11%-46% of clinical cases compared to primary series alone. For severe disease, around 9%-44% more cases could be averted following the fourth dose compared to the primary series alone, across transmission settings (Figure 2). We estimate that a fourth dose delivered 12 months after the primary series would avert roughly one-quarter to one-third more cases than the primary series alone—29%–34% more clinical cases, 27%–31% more severe cases, and 25%–35% more deaths—in settings with *Pf*PR_2-10_ of 20% to 60%. In comparison, a fourth dose delivered 18 months after the primary series would likely avert an additional 11%–12% of clinical cases and 9%–11% of severe cases in the same transmission settings. In lower moderate transmission settings (*Pf*PR =< 10%), while the magnitude of impact is reduced, a fourth dose could still provide between an additional 8%-32% reduction in cases, dependent on dose timing and if sufficient reach is achieved.

We find there is an interaction between dose spacing and the assumptions of the underlying protection against infection that drives value uncertainty in our estimates. We present modelling results for differing protection assumptions with plausible ranges of the fourth dose given at six-, nine-, 12- and 15-months following the primary series and investigate the public health impact. While the impact remains high and is similar if the fourth dose is given six to 12 months after the primary doses, we find different conclusions given a single ideal dose timing. For instance, the nine-month interval (corresponding to 18-months of age at vaccination) could provide the highest cumulative impact as protection is extended to the second year or could be less impactful than a 12-month interval, depending on protection against infection assumptions.

Critically, there is even greater uncertainty around the impact of a 15-month interval (a dose given at 24-months of age). The potential impact of this dose timing is highly dependent on the level of protection against infection of this dose timing (Figures 1 and 2). Specifically, in this study, if the 15-month interval provides the same protection against infection as a 12-month interval then, all else being equal, the slightly longer interval of 15-versus 12-months would lead to a higher cumulative impact over time as a later dose would maintain higher level of protection into the third year of life. However, if the protection against infection is closer to what was observed at the 18-month interval then a dose at 15-months would provide the lowest cumulative impact in most transmission settings. This makes sense considering the trade-off between protection against infection, duration of protection, and the underlying patterns of disease driven by ongoing exposure to *P. falciparum*. A more accurate estimate of impact at this time interval would require further real-world data to better inform the modelled impact estimates.

The proportion of children reached with both the primary series and the fourth dose, measured as coverage, is a crucial driver for impact against malaria morbidity and death. The overall magnitude of the impact, regardless of transmission intensities, is substantially reduced when the reach of the primary series is not sufficiently high, and moderately reduced when the fourth dose reach is limited in the case of a high dropout scenario (Figure 3). We estimate that in a moderate transmission intensity with *Pf*PR_2-10_ of 20%, a third-dose coverage of 50% instead of 80% would result in around 37% reduction in impact on severe malaria cases, while a coverage drop out of 50% from primary series to fourth dose when delivered at a 12-month interval, even with a high third dose reach, would result in around 10% reduction in impact on severe malaria. In an alternate scenario where we assume the paediatric population has much higher probability of receiving timely treatment for uncomplicated malaria at 70%, we found that sufficient reach with the primary series remains the most important driver of impact regardless of transmission setting (Supplemental Figure S8).

**Figure 3.**
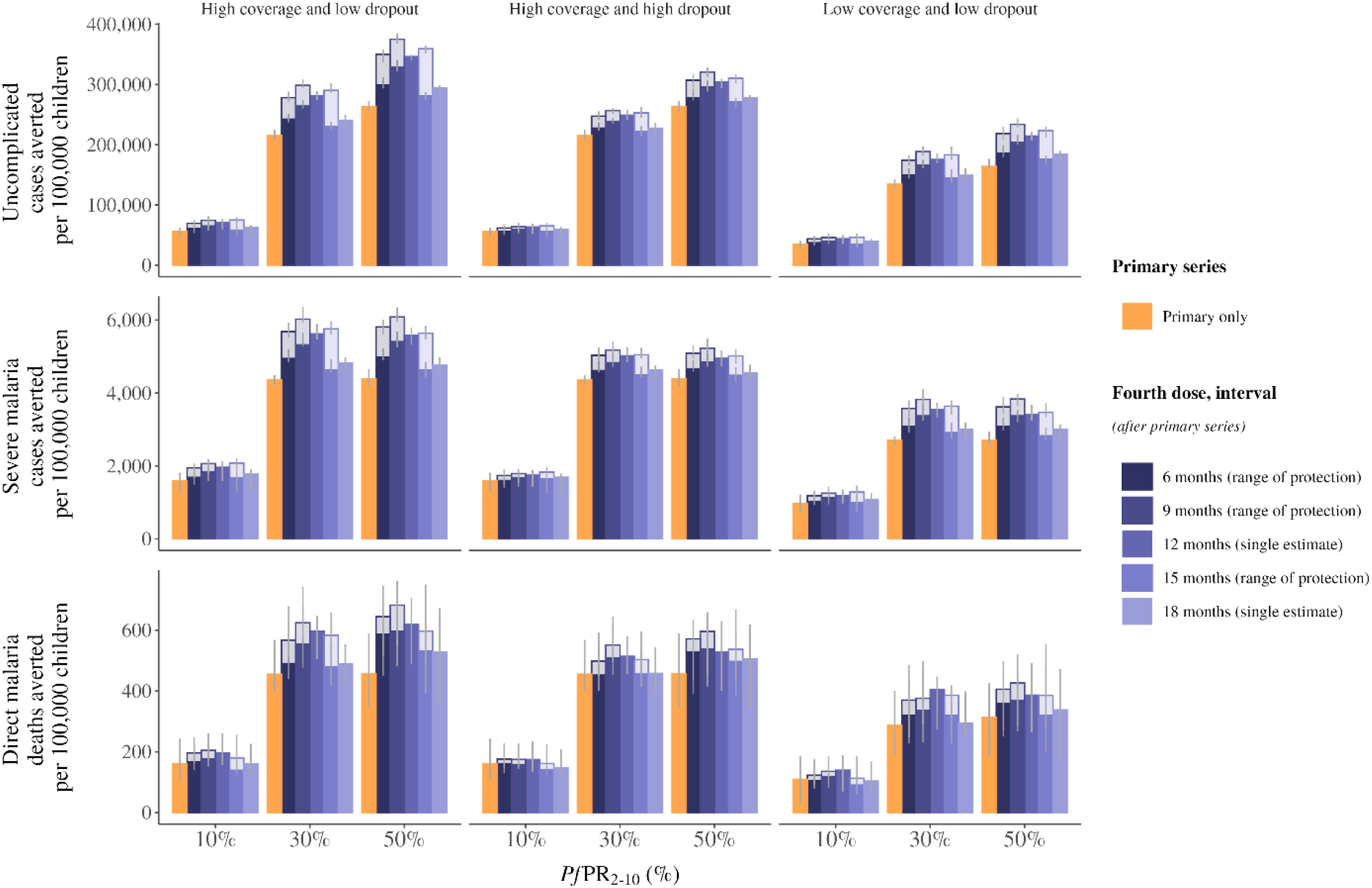
Events averted per 100,000 children under different vaccine coverage rates. The median uncomplicated malaria cases averted (A), severe malaria cases averted (B), and direct malaria deaths (C) averted in children under 5 years of age per 100,000 children comparing the three-dose primary series and a fourth dose delivered at a 6-, 9-, 12-, 15- or 18-month interval after the primary series. Results are shown for three coverage scenarios: high coverage (left) with the primary series coverage of 80% and no dropout between the primary series and fourth dose (fourth dose coverage of 80%); high coverage but high dropout (centre) with the primary series coverage of 80% and a 50% dropout between the primary series and fourth dose (fourth dose coverage of 40%) and; low coverage (right) with the primary series coverage of 50% and no dropout between the primary series (fourth dose coverage of 50%). The two shades on the 6-, 9-, and 15-month intervals represent the range of probable protection against infection values for these three dose timings. Cases averted are calculated over a 10-year time horizon. Error bars show 95% confidence interval across stochastic replicates. Access to care is represented as a 45% 14-day probability to received treatment for uncomplicated malaria.

Finally, it is important to highlight the children who remain unprotected due to being age ineligible (i.e. too young to be vaccinated) in whom a substantial burden of malaria disease and death still remains. The age-specific impact is largely concentrated between nine and 30 months of age (Figure 4 and Supplemental Figures S6-7), with the timing of impact differing by dose timings. Since the burden of symptomatic uncomplicated cases, severe cases and deaths are concentrated in younger children, it stands to reason that a fourth dose of RTS,S vaccine targeting this high-burden age range will have the greatest impact.

**Figure 4.**
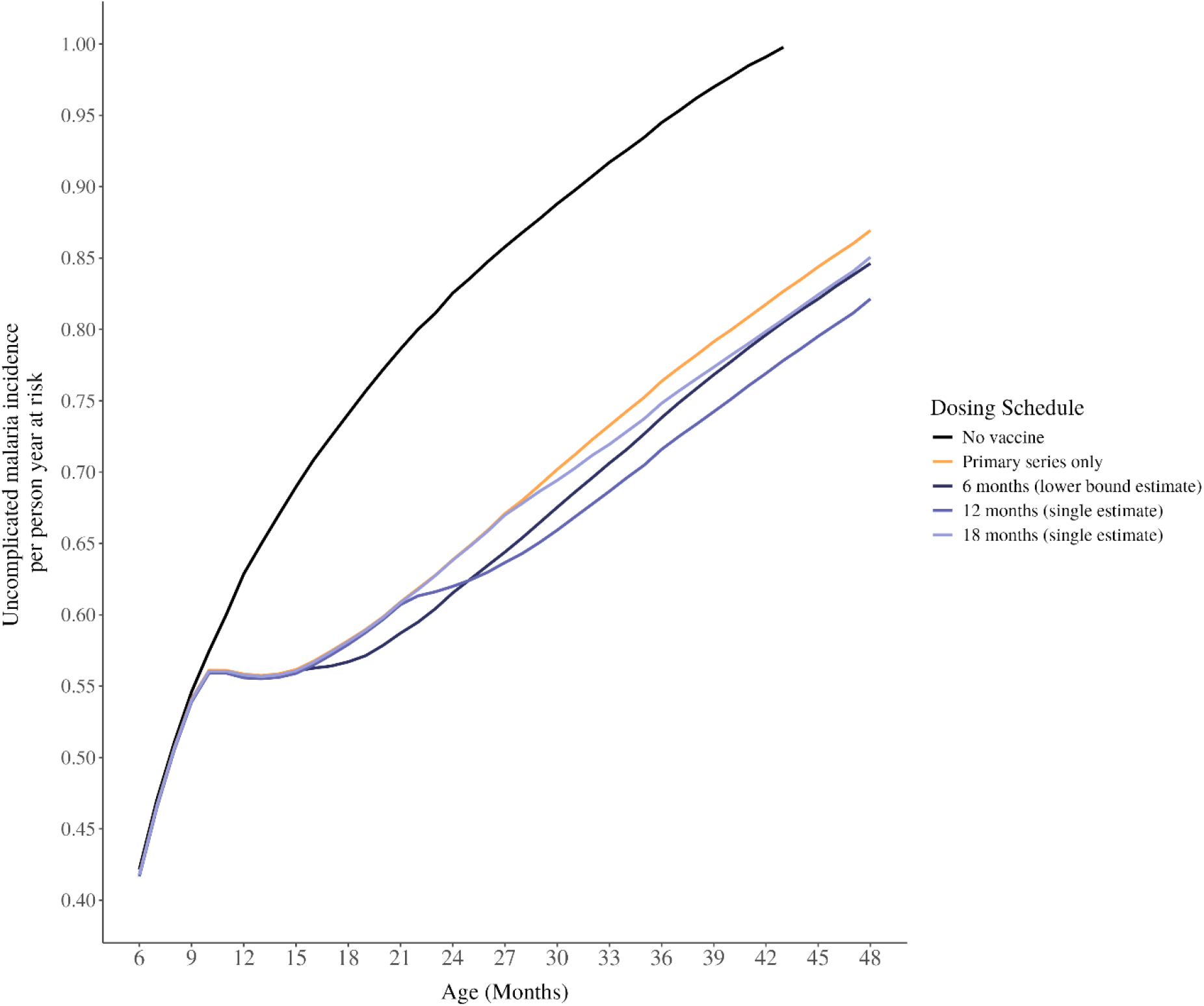
Cumulative incidence rates of uncomplicated malaria cases by age in months, base case. The cumulative incidence rate of uncomplicated malaria cases averted by age in months, over 10 years of follow up. The top panel shows the total differential effects on the age-patterns of uncomplicated disease, and the bottom panel shows a cropped y-axis to better highlight the impact of different dose timings. We show no intervention scenario (black line), three-dose primary series (yellow line), and a fourth dose delivered at a 6-, 9-, 12-, 15- or 18-month interval after the primary series (purple lines). This plot shows the most conservative impact estimates, using the lower bounds of the potential protection against infection for each of the dose timing.

## Discussion

Our modelling results show that RTS,S malaria vaccine impact largely depends on the number of children reached with a vaccine for both the three-dose primary series and the fourth dose. To maximize lives saved, achieving high coverage of the primary series should remain a top priority, alongside efforts to ensure the fourth dose is accessed. When a high proportion of targeted children are vaccinated, our analysis confirm that a fourth dose deployed between six to 12 months after the primary series leads to the highest burden averted. Hence, the fourth dose should be timed to prioritize touchpoints that will maximize coverage. While the absolute magnitude of impact may vary, these conclusions remain across transmission intensities. Overall, our results support the flexible timing of a fourth dose of RTS,S malaria vaccine to align with an existing immunization touchpoint in the second year of life to maximize coverage and optimize the impact in perennial settings.

A recent modelling study by Thompson et al. looking at dose timing in seasonal settings came to a similar final conclusion around potential flexibility in dose timing, with the aim of improving access to the vaccine in seasonal settings (24). These conclusions align with the understanding the age-distribution of severe disease and death, where the burden remains highly concentrated in young children under 2 years of age (35,36). Critically, the current vaccination schedule for malaria vaccines continues to leave an important portion of children unprotected due to being too young for the current vaccine (i.e. less than five or six months old at the start of the primary series) (Figure 4).

Our results for the absolute burden averted as shown for the different coverage scenarios reinforce the crucial importance of the proportion of children reached with the vaccine. While we estimate different magnitudes of impact based on incrementally different dose timings and plausible protection against infection, we did not find any scenario where a high vaccine protection against infection and optimized dose timing under low coverage assumptions outperformed a higher coverage scenario. Ultimately, reaching a sufficiently large number of children with the RTS,S vaccine by leveraging existing systems should be prioritized to achieve desired impact. This will, hopefully, allow for flexibility in the timing of the fourth dose to align to country-specific schedules and needs.

The protection against infection for six-, nine- and 15-month intervals are presented as a plausible range broadly showing that the vaccine would ideally be delivered at the latest possible interval with the highest protection. However, for the simplicity of the simulations, we only simulated the extremes of the plausible ranges based on existing evidence. We know, based on immunological data from the Phase 3 trial (8) and previous calibration (11) that the fourth dose provides some return to protection; however, we do not know to what level and how both the fourth dose interval after the third as well as the age of vaccination effect this “boosting” effect.

Particularly challenging was defining a plausible range for the protection against infection of the fourth dose for the 15-month interval. This interval corresponds to a dose given at 24-months of age, which is of high interest to many countries as it could contribute to strengthening a second-year of life platform alongside other booster and catch-up vaccines (37). Unfortunately, the bounds on the assumptions of protection against infection are high due to not having data to inform vaccine assumptions, and therefore, this dose could potentially provide the most or the least cumulative impact depending on the true protection against infection of this dose timing. Because of the wide plausible efficacy range at this interval, countries considering a fourth dose at 24-months of age may benefit most from additional real-world data on this dose timing.

As with all modelling studies, there are several limitations in our study including necessary simplification of modelling assumption and insufficient data to inform protection assumptions of vaccine timings outside existing evidence. Firstly, our study aimed to provide additional evidence to inform global policy setting by exploring multiple scenarios across a range of malaria transmissions and levels of access to case management for symptomatic cases to identify any important trends or signals. Therefore, we model a broad set of archetypal settings and assume a fixed level of access to care, monthly and annual transmission, and population-level access to the vaccine. This modelling is not meant to recreate specific country scenarios, which would be best carried out by or in close collaboration with the individual country; instead, it sets to define the broad bounds within which a global policy decision can be taken.

Secondly, our model estimates rely on assumptions around the initial protection against infection at different dose timings. However, data to inform these parameters are only available for the primary series (11,13), 12-month interval (33), and 18-month interval (11,13). These results are aligned with results from other models regarding original calibration to the Phase 3 trials (13), and updated calibration of models to newer real-world implementation data (24). However, there is no evidence-based estimates for vaccine protection against infection at intervals shorter than 12-months post-primary series, nor for the 15-month interval. To mitigate the limitations around the protection against infection assumption, we present a range of plausible values around these implementation scenarios to explore the possible public health impact. Still, based on our model structure and the available evidence, we are unable to assume any differences in immunological response for shorter time intervals, which could affect our impact assessments. These estimates should be updated once additional evidence on alternate dose timings become available.

Lastly, we use OpenMalaria, an established and validated individual-based model of malaria transmission and intervention dynamics for this study. However, as with all models there are structural limitations. Relevant to this study are model parameters that determine age-patterns of severe disease and deaths that were calibrated to older data of severe disease, while we know patterns of underlying disease and co-morbidities may have changed over time. Nevertheless, a recent comparison of our model with contemporary data on severe disease from Kenya Health and Demographic Surveillance Sites (HDSS) (38) shows that the model is still able to recover severe malaria trends seen in contemporary real-world data.

These considerations of timing of additional doses of another vaccine, or injectable, are relevant beyond the RTS,S/AS01 context. There are other pre-erythrocytic vaccines in Phase II and Phase III trials, as well as a steady pipeline of forthcoming vaccines and injectable products (39–41). As the second licensed malaria vaccine, R21/Matrix-M is also introduced across the sub-continent, similar questions have already arisen around optimal deployment of the vaccine. While modelling based on immune correlates has shown a different protection profile than RTS,S vaccine (42), expanding the reach of this vaccine will likely remain the most important determinant of population-level impact. The effective and impactful deployment of any new intervention will depend on finding the delicate balance between optimized immunological benefit and realistic delivery strategies, to maximize real world.

## Conclusions

To maximize the public health impact of the RTS,S malaria vaccine, countries should prioritize achieving high coverage of the three-dose primary series, followed by a flexibly timed fourth dose to ensure higher coverage of this fourth dose. Our modelling in perennial settings shows that vaccine impact is primarily driven by the number of children reached—regardless of the exact timing of the fourth dose. When high coverage is achieved, delivering a fourth dose between 6 and 12 months after the third dose consistently results in the greatest burden averted. These findings hold across transmission intensities, reinforcing that the fourth dose should be aligned with existing immunisation touchpoints within the 15– 24-month age range that can maximize uptake. If strong touch points do not exist, it would be prudent to strengthen those within an existing system to increase access to all doses of malaria vaccine. As real-world rollout of malaria vaccines that include RTS,S/AS01 and R21/Matrix-M expands, evaluating the effectiveness of flexible fourth dose strategies will be key to informing national implementation and optimising long-term malaria control.

## Data Availability

All data produced are available online at

https://doi.org/10.5281/zenodo.17342794

## Availability of data and materials

Model inputs and scripts supporting the conclusions of this article are available at https://doi.org/10.5281/zenodo.17342794.

## Acknowledgements

We thank Mary Hamel, Eliane Furrer, Scott Gordon, Saira Nawaz, and Hayley Thompson for discussions following these analyses. In addition, we acknowledge all members of the Disease Modelling research unit of the Swiss Tropical and Public Health Institute and the Global Disease Modelling team at The Kids Research Institute for their advice. Calculations were performed at sciCORE (http://scicore.unibas.ch/) scientific computing center at University of Basel.

## Funding

This study was funded by the Gates Foundation (INV-025569 to MAP).

MAP acknowledges support from the Swiss National Science Foundation (SNF Professorship PP00P3_203450 to MAP). The funders had no role in the design or analysis of this study.

### Contributions

DFD and MAP conceived the study. MAP provided overall guidance while DFD, JM and AC were involved in developing the modelling framework. DFD and JM conducted the analysis. DFD and JM analysed results and prepared figures. SLK and MAP supported the interpretation of results. DFD wrote the first draft of the manuscript. All authors provided input and critically reviewed the manuscript and approved the final draft for submission.

### Corresponding Author

Correspondence to Melissa A. Penny.

## Ethics declarations

### Ethics approval and consent to participate

Not applicable.

### Consent for publication

Not applicable.

### Completing interests

The authors declare no competing interests.

## Notes

### Competing Interest Statement

The authors have declared no competing interest.

