## Supplement 1 for "Timing of the fourth dose of RTS,S/AS01 malaria vaccine in perennial settings: a modelling study"

### Methods

#### Mathematical Transmission Model

OpenMalaria is an individual-based, stochastic model of *Plasmodium falciparum* malaria epidemiology in humans linked with a deterministic model of mosquito dynamics and infectiousness. The model includes components of blood-stage paradise densities, dynamics of infection in humans, human health dynamics and incidence of co-morbidities. Details on the model structure, calibration and historical fitting have been previously published (1–3). All model variants in OpenMalaria have been fit to real-world data from malaria endemic areas, as described in previous published literature. **Table S1** provides a summary of the key core assumptions of the model. Additionally, the open-source code is available on (https://github.com/SwissTPH/openmalaria/wiki), including detailed documentation.

The model attempts to capture the dynamics during the whole Plasmodium parasite lifecycle through both mosquito and human hosts. Critically, the model uses an entomological inoculation rate (EIR) which drives the force of infection in humans. This subsequently translates into probabilities of blood stage and liver stage infection, progression to detectable infection (by RDT or PCR for example), presentation with clinical, uncomplicated malaria, and progression to severe malaria, hospitalization with or without sequelae and/or death in the human host.

**Table S1. Summary of the OpenMalaria model components adapted from Golombeau et al., Galactionova et al., and Reiker et al.** (1,3,4)

| Component | Description of core assumption |
|---|---|
| *Key modelled epidemiological processes (base model)* | |
| Malaria infection of humans<br><br>(5) and eq. 1-4 of Additional file 1 in (6) | • Determined by EIR which is a model input and affects the force of infection in the simulated setting - Considers an age-dependent exposure of human hosts to mosquitoes (correlating with body-surface area)<br>• The relationship between infection rates and EIR is defined and fitted with data from The Gambia, Nigeria and Kenya in (5) |
| Infection progression in humans: asexual parasite densities and immunity<br><br>(5–8) and eq. 5-15 of Additional file 1 in (6) | • Blood-stage parasite density depends on the time since infection and is affected by naturally acquired immunity. Acquired immunity reduces parasite density of subsequent infections.<br>• The duration of infection follows a log-normal distribution and is estimated from a malaria therapy dataset ((8) and eq. 1 in (7))<br>• Immunity (both pre-erythrocytic and blood-stage) develops progressively following consequent episodes of exposure to infection and total parasitaemia seen by an individual in their lifetime.<br>• Super-infection is possible with cumulative parasite densities - The parasite density in a host at a given time is defined and fitted with data from Ghana, Nigeria and Tanzania in (7) |
| Transmission from infected humans to mosquitoes (6,9,10) and eq. 16-21 of Additional file 1 in (6) | • Infectivity to mosquitoes depends on the density of parasites present in the human (including a time-lag for gametocyte development)<br>• The fraction of resulting infected mosquitoes after feeding on a human host follows a binomial distribution - The relationship between infectivity to mosquitoes and parasite density was defined and fitted in (9) with data from malaria therapy collected in Georgia between 1940 and 1963 and available from (8)<br>• The age-specific contribution to overall infectiousness to mosquitoes was validated in (9) against field data collected from Liberia, The Gambia, Tanzania, Kenya, Papua New Guinea and Cameroon. |
| Clinical illness, morbidity, mortality, and anaemia (2,6,11,12) and eq. 22-32 of Additional file 1 in (6) | • Acute clinical illness depends on human host parasite densities and their pyrogenic threshold which evolves over time depending on the individual exposure history<br>• Acute morbidity episodes can be uncomplicated or evolve to severe episodes; a proportion of the severe episodes leads to deaths<br>• The probability of a clinical malaria episode was defined and fitted with data from Senegal in (11)<br>• The probabilities that a clinical episode becomes severe and the risk of mortality for a severe episode are defined and fitted to field data from over 10 African countries in (2) |
| *Modelled characteristics of the transmission setting* | |
| Population age structure (2,13) | • Informed by health and demographic surveillance data from Tanzania |
| Transmission seasonality (5,14) | • Seasonally forced, the same transmission pattern is reproduced each year in absence of interventions, displayed in Fig. S2.1. Seasonal patterns are inputs to the model and users can define any patterns as needed e.g., perennial, one or two peak seasonal patterns, etc. |
| Case management (15) | • Modelled through a comprehensive decision tree-based model defined and validated in (15) which determines the corresponding treatment implications depending on the occurring clinical events such as fevers and seeking of care<br>• Its representation includes specification of access to official or nonofficial care, access to hospital for severe cases, diagnostic tests (use, specificity, sensitivity, and threshold of detection), treatments for first, second line and non-official care, effects of treatment, case fatality rate, case sequelae and cure rates. |
| Entomological setting (16) | • Comprehensive simulation of the mosquito lifecycle and behaviour towards human and animal hosts (biting, resting) embedded in a |

| | |
|---|---|
| | dynamic entomological model of the mosquito feeding cycle defined in (16)<br>• Multiple vector species can be simulated simultaneously Modelled interventions and their action Vector control (17–19)<br>• Acts on the availability of the protected human hosts to mosquitoes (deterrence), on the probability that a mosquito bites a protected human host (pre-prandial effect) and on the probability that a mosquito survives host feeding (postprandial effect) |
| Drugs and Vaccines (20,21) | • Act at various levels of the parasite life cycle (transmission blocking, anti-infective, blood-stage clearance) and their action is defined by their initial protection against infection, half-life, and decay |
| RTS,S vaccine (22,23) | Parameterized against RTS,S Phase 3 clinical trial data (24) and subsequently against RTS,S vaccine delivery in real-world settings (23).<br>• Decay: adapted Weibull distribution with a decay shape of 0.69 (22), characterized by rapid initial decay followed by plateau<br>• Half life: 7.32 months (22)<br>• Initial protection against infection of primary series: 91% (22)<br>• Initial protection against infection of fourth dose delivered 12-months after the primary series: 77% (23)<br>• Initial protection against infection of fourth dose delivered 18-months after the primary series: 49% (22) |
| Deployment characteristics | • Interventions can be deployed for several rounds to a targeted group of individuals and specified coverages (proportion of the population covered by the intervention) |
| *Simulation regimes and model variants* | |
| Time steps | • Simulation outputs are tracked every 5 days |
| Model variants (25) | • Varying assumptions in immunity decay, treatment and heterogeneity of transmission are covered in 14 model variants described in (25). In the present study, we use the base model (parameterization described in Table 2 in (25) under the denomination R0001) |
| *Software availability and documentation* | |
| Source code and wiki page available on GitHub: https://github.com/SwissTPH/openmalaria/ | |

### RTS,S vaccine parameters

In OpenMalaria, the deployment of a pre-erythrocytic vaccine like RTS,S/AS01 is modelled as the prevention of sporozoites or a reduction in the probability of infection. The protection dynamics profile of the vaccine is assumed to follow an adapted Weibull distribution in the form of $\cdot\ e^{-(t/b)^{c} \cdot \log(2)}$, where $a$ is an initial protection against infection, $b$ is the half-life of protection and $c$ represents the Weibull decay shape. This profile assumes a rapid initial decay followed by a second, less rapid phase of decay. Notably, we assume the same half-life and decay pattern for all the fourth doses, regardless of timing.

OpenMalaria was calibrated to the Phase 3 clinical trial data for RTS,S (5) to derive the underlying properties of the vaccine, including an initial protection against infection parameter following the primary series, and a protection against infection parameter following a fourth dose delivered 18-months following the primary series. In 2024, Malinga et al. (23) fit this same model to data on RTS,S vaccine given with and without seasonal malaria chemoprevention (SMC) (26,27) in seasonal settings to estimate a parameter of protection against infection of a fourth dose at a 12-month dosing interval. **Figure S1** shows a graphical representation of the efficacy profile assumptions, combining the protection against infection and the decay assumptions.

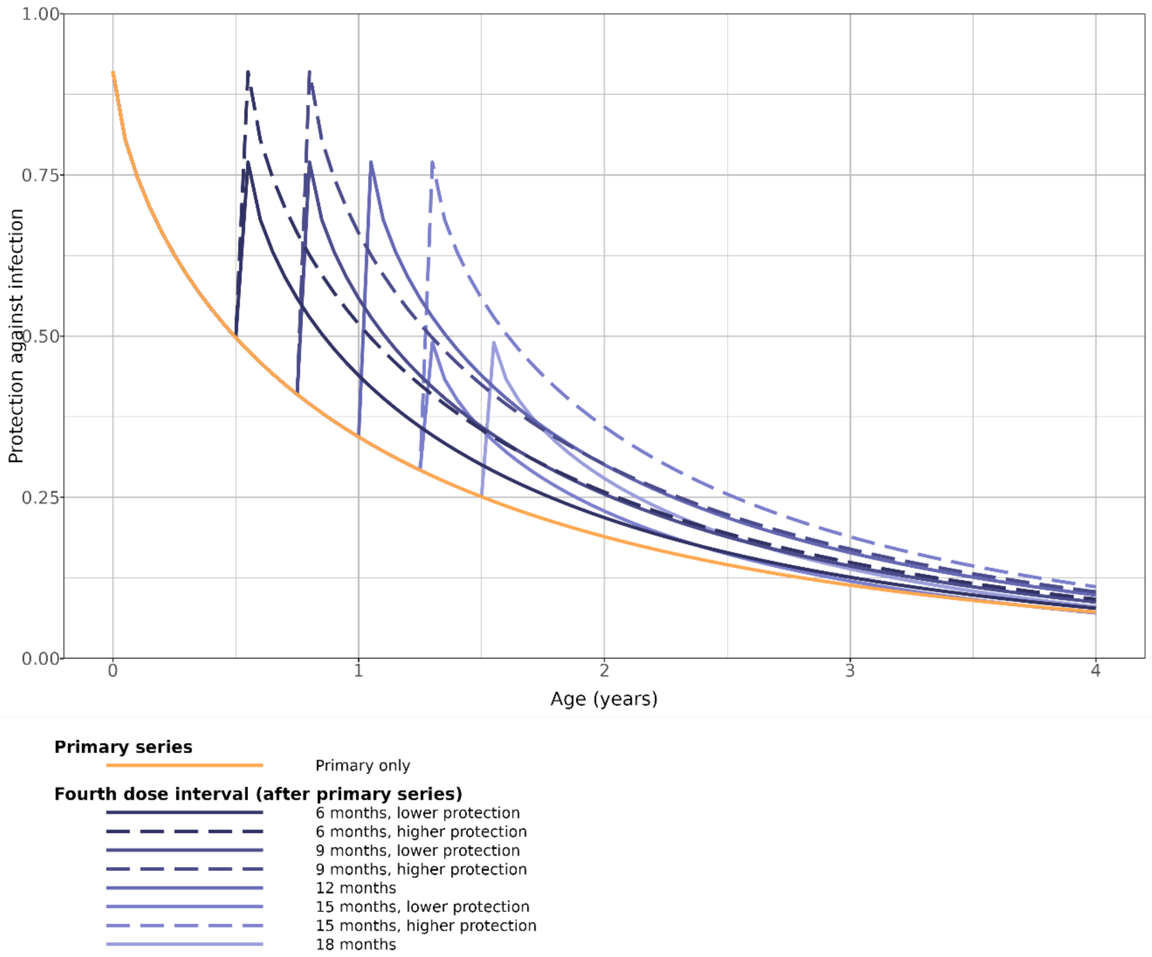


***Figure S1.* Underlying assumptions of protection against infection comparing of the different dose timings.**

*Protection against infection of the three-dose primary series (yellow line) and a fourth dose delivered at a 6-, 9-, 12-, 15- or 18-month interval after the primary series (purple lines). These lines represent three model parameters: initial protection against infection (shows as the peak), half-life, and decay rate. The dashed lines at 6-, 9-, and 15-month intervals represent the upper range of probable protection against infection values for these three dose timings.*

The OpenMalaria RTS,S vaccine parameterization has been validated against vaccine efficacy estimates from both the 7-year follow up of the Phase 3 clinical trial data as well as the 46-month follow up data from the Malaria Vaccine Implementation Pilots (MVIP). The model has also been reviewed by WHO SAGE/IVIR-AC as part of their consideration of modelling data for policy setting.

### **Disease definitions** (20)

Infection with *Plasmodium falciparum* can lead to a number of different health outcomes, although the gradual acquisition of immunity over time means that symptomatic cases tend to be more prominent in younger age groups while older ages tend do experience more sporadic episodes and high rates of asymptomatic carriage (28). Young children are particularly at risk of severe disease, which is a term for life-threating forms of malaria mainly driven by severe anaemia or cerebral malaria (29,30). There is no single definition of severe disease but in general this form of disease requires hospitalisation. Clinical, or uncomplicated, malaria generally refers to the less severe form of malaria measured as fever plus parasite density in the blood over some threshold, although again there is no single standard definition.

For this study, we calculated the following outcomes:

Clinical cases, or uncomplicated malaria. In OpenMalaria, multiple episodes of clinical cases can occur per infection.

Severe cases, which is directly predicted from OpenMalaria, including those receiving in-patient treatment and those that would be classified as severe in the community.

Mortality, or deaths, which include (a) direct deaths that are directly attributable to malaria, and (b) all-malaria deaths, which include deaths resulting from malaria in the presence of a co-incurring morbidity.

### Description of Outcomes

We examine a range of transmission intensities, defined by the mean *Plasmodium falciparum* parasite prevalence in children aged between two and ten years (*Pf*$PR_{2\text{-}10}$), detectable by rapid diagnostic test. We measure certain outcomes as cases averted per 100,000 fully vaccinated children where a fully vaccinated child is defined as a child who received the three-dose vaccine primary series.

### Supplementary Results

#### *Impact on indirect deaths under high coverage assumptions*

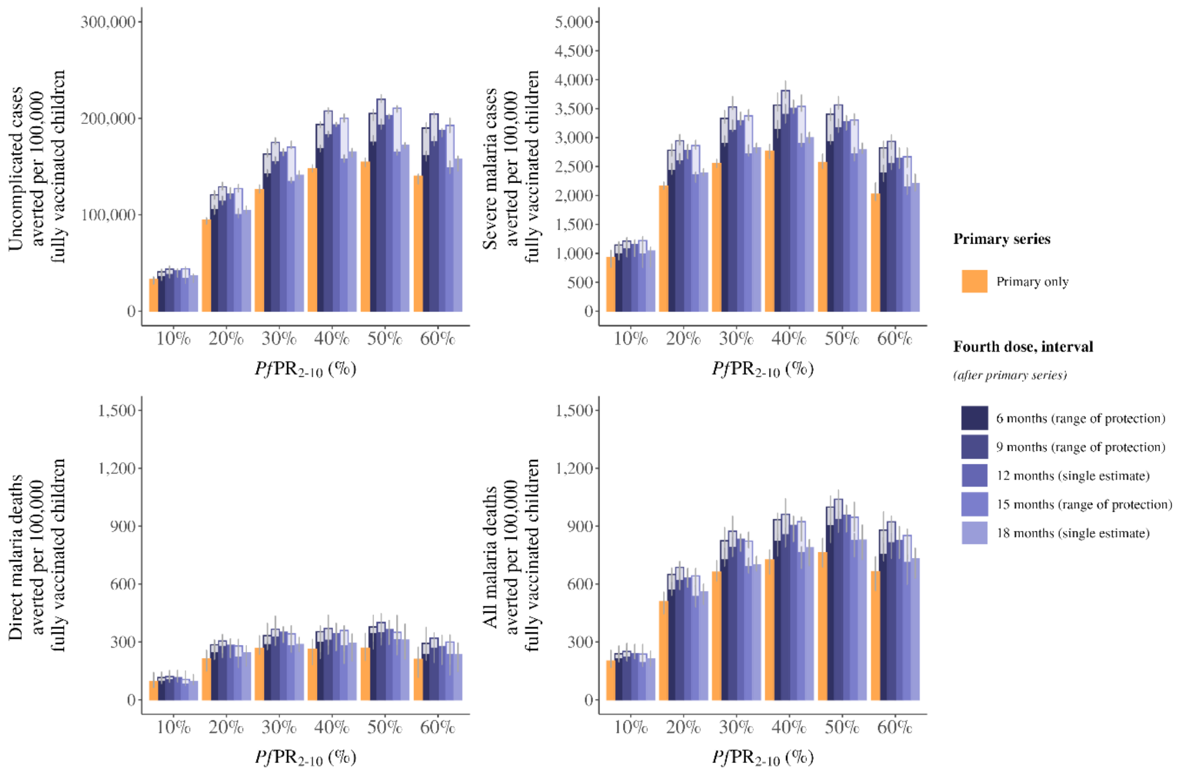


***Figure S2 . Events averted per 100,000 fully vaccinated children, comparing timing of fourth dose of RTS,S malaria vaccine.***

*The median uncomplicated malaria cases averted (A), severe malaria cases averted (B), direct malaria deaths averted (C) and all malaria deaths averted (D) in children under 5 years of age per 100,000 fully vaccinated children comparing the three-dose primary series and a fourth dose delivered at a 6-, 9-, 12-, 15- or 18-month interval after the primary series. The two shades on the 6-, 9-, and 15-month intervals represent the range of probable protection against infection values for these three dose timings.  Cases averted are calculated over a 10-year time horizon. This is a default reference scenario of high coverage, where primary series coverage is assumed at 80% of the target population and there is no drop-out between the primary series and the fourth dose. Error bars show 95% confidence interval across stochastic replicates. Access to care is represented as a 45% 14-day probability to received treatment for uncomplicated malaria.*

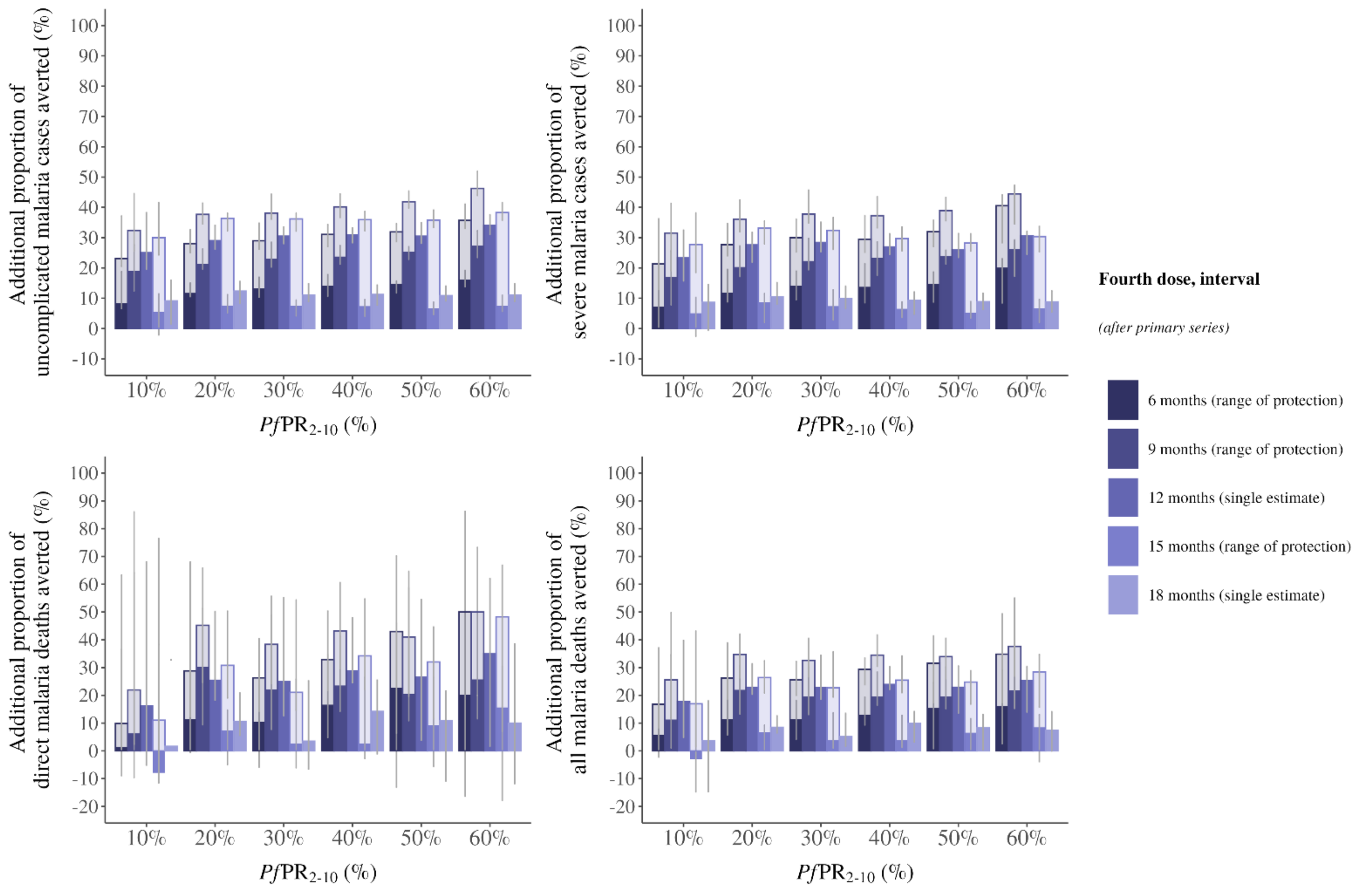


***Figure S3. Proportion of additional cases averted by a fourth dose of RTS,S malaria vaccine.***

*The proportion of additional uncomplicated malaria cases averted (A), severe malaria cases averted (B), direct malaria deaths averted (C) and all malaria deaths averted (D) in children under 5 years of age per 100,000 fully vaccinated children at different timings of a fourth dose delivered at a 6-, 9-, 12-, 15- or 18-month interval after the primary series, compared to the primary series alone. The two shades on the 6-, 9-, and 15-month intervals represent the range of probable protection against infection values for these three dose timings. Cases averted are calculated over a 10-year time horizon. This is a default reference scenario of high coverage, where primary series coverage is assumed at 80% of the target population and there is no drop-out between the primary series and the fourth dose. Error bars show 95% confidence interval across stochastic replicates. Access to care is represented as a 45% 14-day probability to received treatment for uncomplicated malaria.*

### Impact under different coverage assumptions

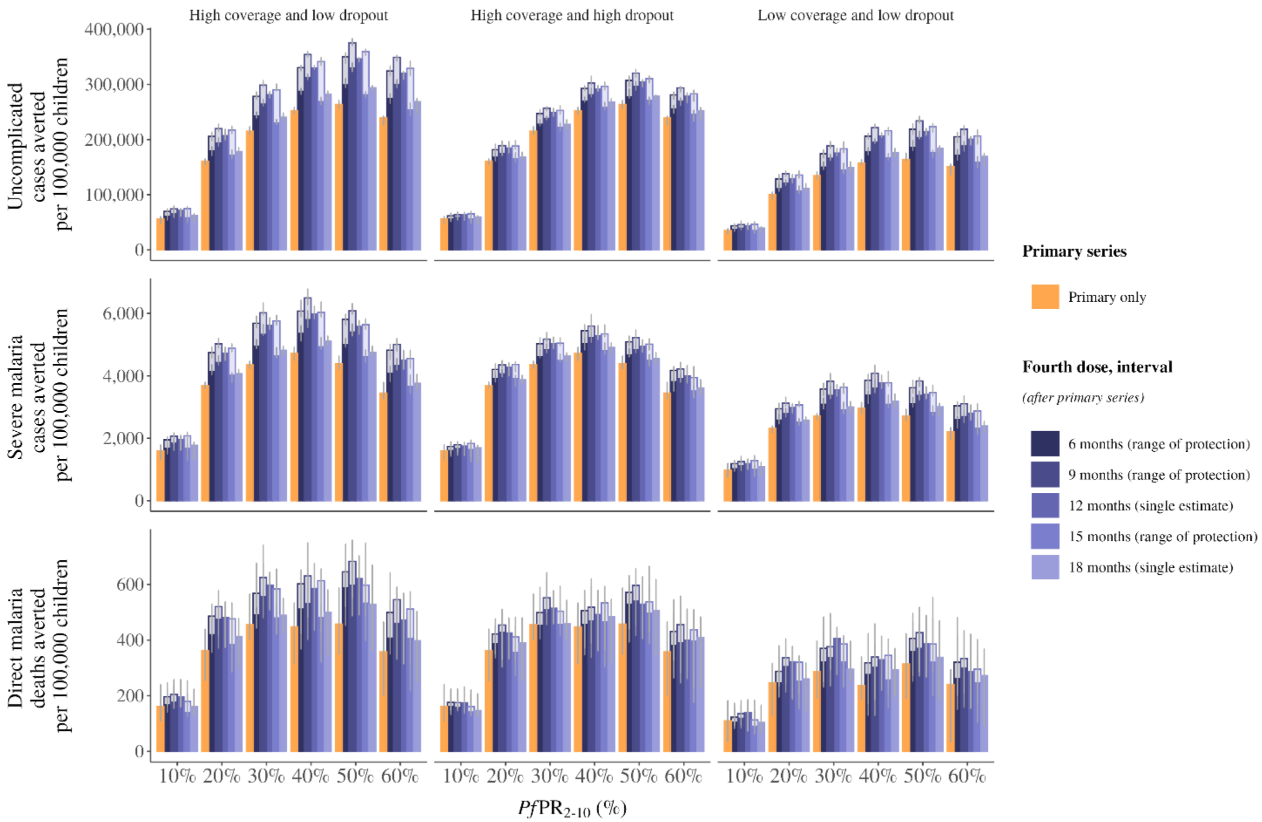


***Figure S4. Events averted per 100,000 children under different vaccine coverage rates.***

*The median uncomplicated malaria cases averted (A), severe malaria cases averted (B), and direct malaria deaths (C) averted in children under 5 years of age per 100,000 children comparing the three-dose primary series and a fourth dose delivered at a 6-, 9-, 12-, 15- or 18-month interval after the primary series. Results are shown for three coverage scenarios: high coverage (left) with the primary series coverage of 80% and no dropout between the primary series and fourth dose (fourth dose coverage of 80%); high coverage but high dropout (centre) with the primary series coverage of 80% and a 50% dropout between the primary series and fourth dose (fourth dose coverage of 40%) and; low coverage (right) with the primary series coverage of 50% and no dropout between the primary series (fourth dose coverage of 50%). The two shades on the 6-, 9-, and 15-month intervals represent the range of probable protection against infection values for these three dose timings. Cases averted are calculated over a 10-year time horizon. Error bars show 95% confidence interval across stochastic replicates. Access to care is represented as a 45% 14-day probability to received treatment for uncomplicated malaria.*

### *Additional cases averted with low fourth dose coverage (high dropout)*

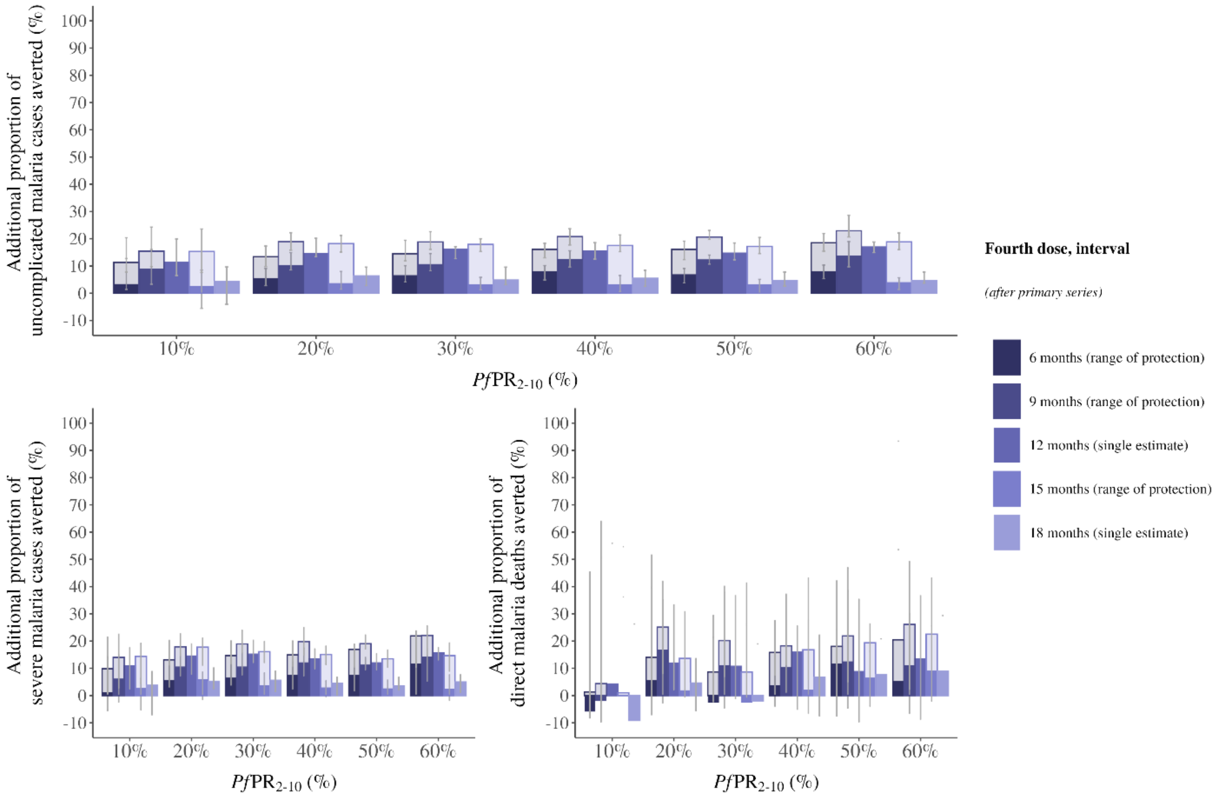


**Figure S5. Proportion of additional cases averted by a fourth dose of RTS,S malaria vaccine.**

*The proportion of additional uncomplicated malaria cases averted (A), severe malaria cases averted (B), direct malaria deaths averted (C) and all malaria deaths averted (D) in children under 5 years of age per 100,000 fully vaccinated children at different timings of a fourth dose delivered at a 6-, 9-, 12-, 15- or 18-month interval after the primary series, compared to the primary series alone. The two shades on the 6-, 9-, and 15-month intervals represent the range of probable protection against infection values for these three dose timings. Cases averted are calculated over a 10-year time horizon. This is a default reference scenario of high coverage but high dropout (centre) with the primary series coverage of 80% and a 50% dropout between the primary series and fourth dose (fourth dose coverage of 40%). Error bars show 95% confidence interval across stochastic replicates. Access to care is represented as a 45% 14-day probability to received treatment for uncomplicated malaria.*

### *Age patterns of disease*

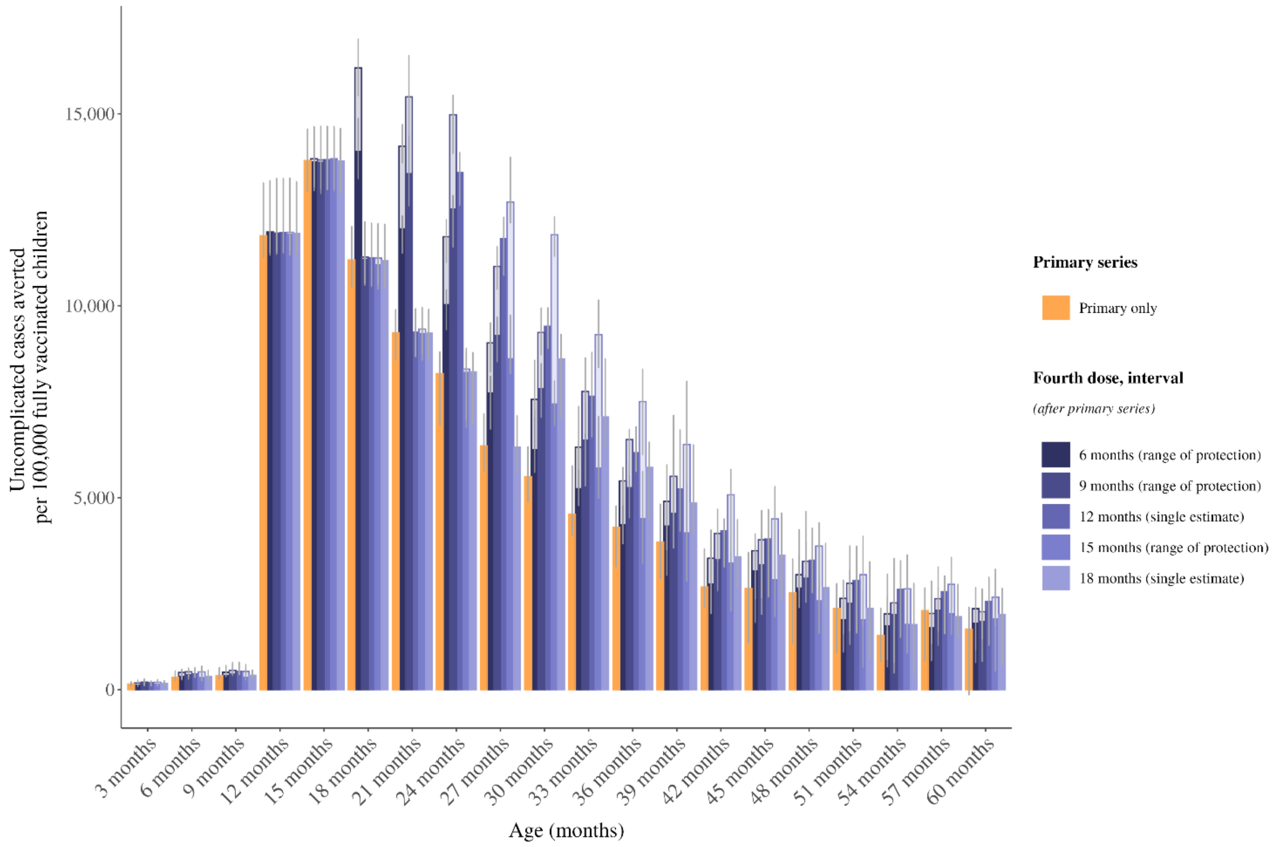


***Figure S6. Uncomplicated malaria averted by age comparing scenarios in a moderate transmission setting with PfPR$_{2\text{-}10}$ of 20%.***

*The cumulative number of uncomplicated malaria cases averted by age in months, over 10 years of follow up. We show three-dose primary series (yellow lines), and a fourth dose delivered at a 6-, 9-, 12-, 15- or 18-month interval after the primary series (purple lines) compared to a no intervention baseline. The two shades on the 6-, 9-, and 15-month intervals represent the range of probable protection against infection values for these three dose timings. Cases averted are calculated over a 10-year time horizon. Error bars show 95% confidence interval across stochastic replicates. Access to care is represented as a 45% 14-day probability to received treatment for uncomplicated malaria.*

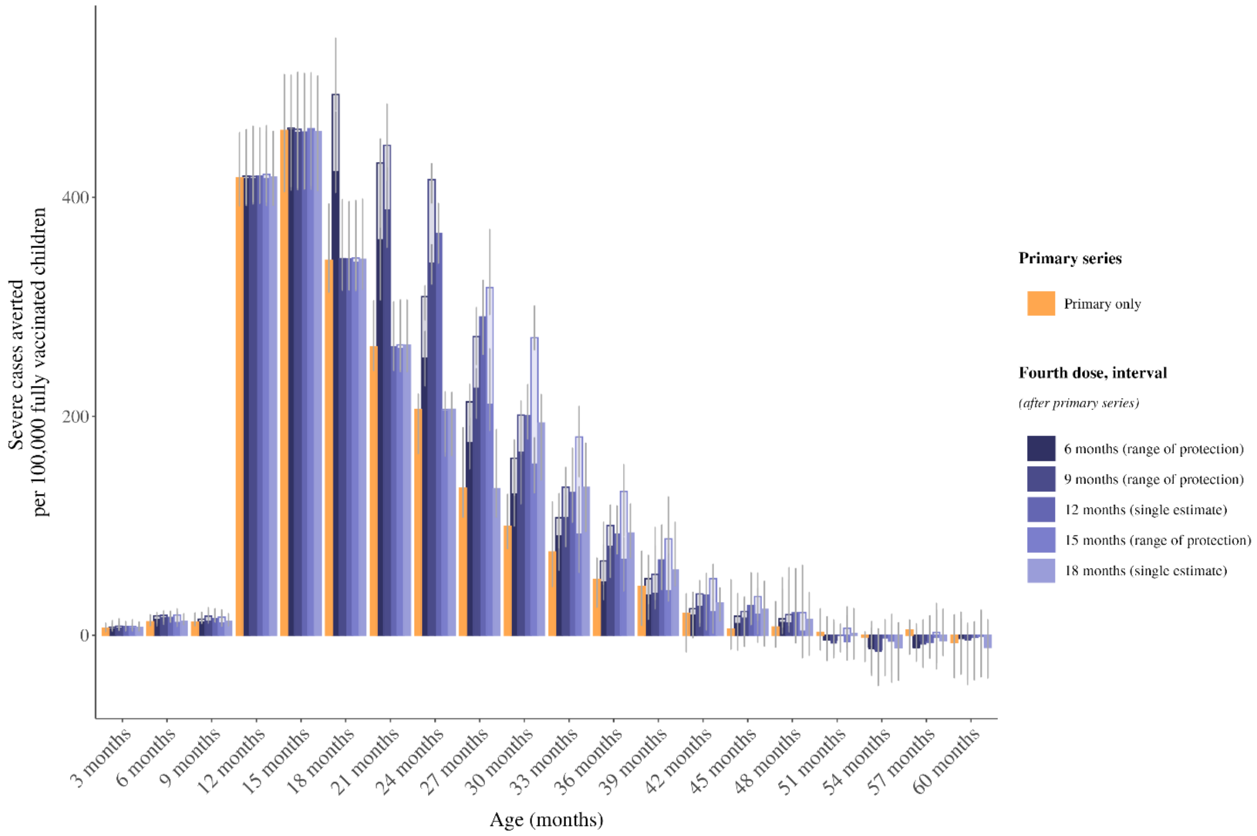


***Figure S7. Severe malaria averted by age comparing scenarios in a moderate transmission setting with $PfPR_{2\text{-}10}$ of 20%.***

*The cumulative number of severe malaria cases averted by age in months, over 10 years of follow up. We show three-dose primary series (yellow lines), and a fourth dose delivered at a 6-, 9-, 12-, 15- or 18-month interval after the primary series (purple lines) compared to a no intervention baseline. The two shades on the 6-, 9-, and 15-month intervals represent the range of probable protection against infection values for these three dose timings. Cases averted are calculated over a 10-year time horizon. Error bars show 95% confidence interval across stochastic replicates. Access to care is represented as a 45% 14-day probability to received treatment for uncomplicated malaria.*

### Impact under high access to care assumptions

The default reference scenario presented in this analysis assumes a 14-day probability to receive treatment for uncomplicated malaria of 45%. As a supplementary analysis, we conducted the same analysis as outlined in Table 1; however, we assume a high 14-day probability to receive treatment for uncomplicated malaria at 70%.

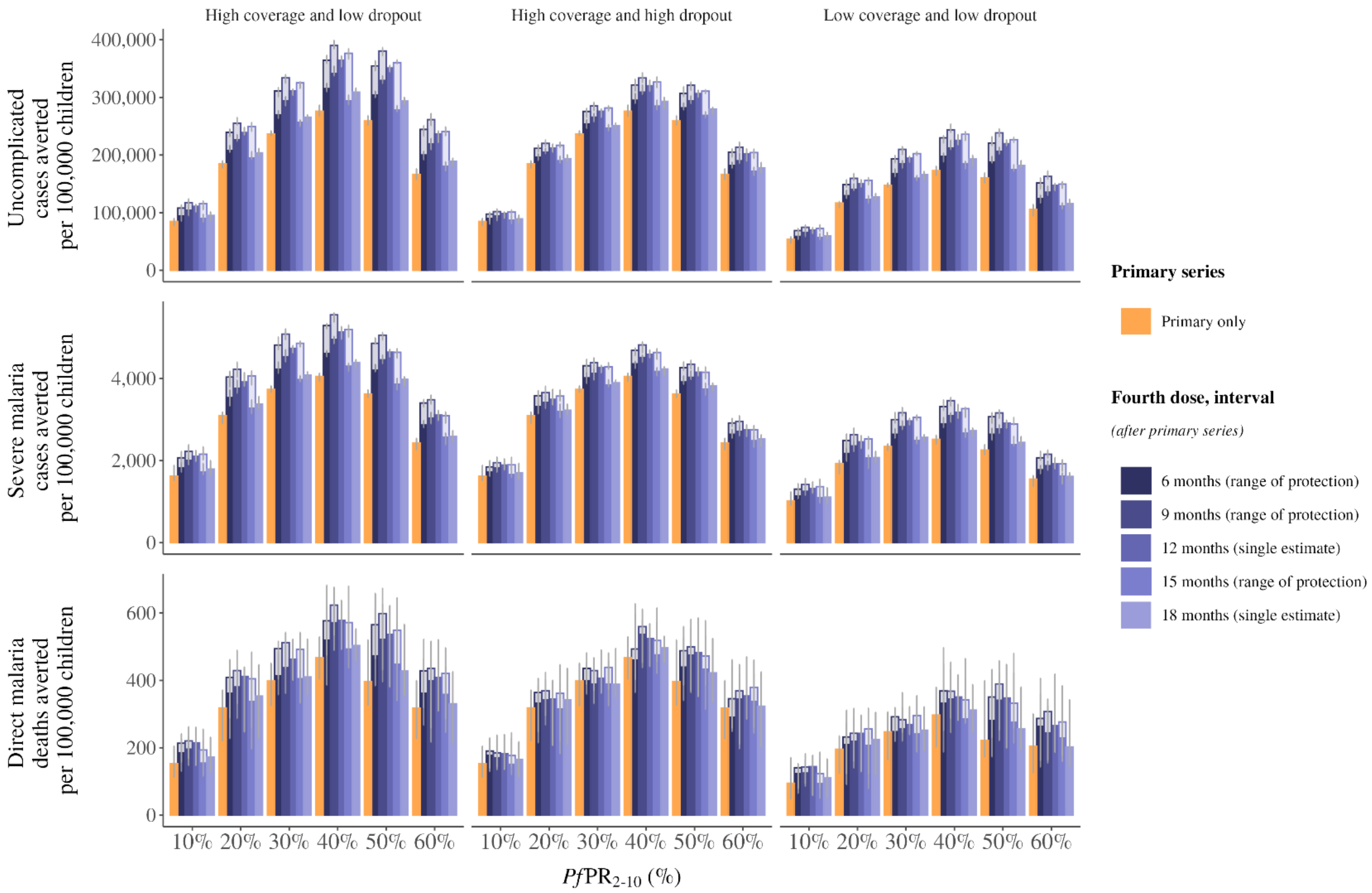


***Figure S8. Events averted per 100,000 children under different vaccine coverage rates.***

*The median uncomplicated malaria cases averted (A), severe malaria cases averted (B), and direct malaria deaths (C) averted in children under 5 years of age per 100,000 children comparing the three-dose primary series and a fourth dose delivered at a 6-, 9-, 12-, 15- or 18-month interval after the primary series. Results are shown for three coverage scenarios: high coverage (left) with the primary series coverage of 80% and no dropout between the primary series and fourth dose (fourth dose coverage of 80%); high coverage but high dropout (centre) with the primary series coverage of 80% and a 50% dropout between the primary series and fourth dose (fourth dose coverage of 40%) and; low coverage (right) with the primary series coverage of 50% and no dropout between the primary series (fourth dose coverage of 50%). The two shades on the 6-, 9-, and 15-month intervals represent the range of probable protection against infection values for these three dose timings.*

30. World malaria report 2024: addressing inequity in the global malaria response. Geneva: World Health Organization; 2024.
